# Comparable Outcomes for Exclusive Lipofilling and Lipofilling-Implant Combination in Breast Reconstruction: A Retrospective Cohort Study

**DOI:** 10.1101/2023.06.13.23291323

**Authors:** Kais Razzouk, Alfred Fitoussi, Arash Rafii Tabrizi

## Abstract

**Background:** Autologous fat grafting combined with prosthesis placement has become a widely accepted standard for breast reconstruction. However, a growing trend toward exclusive lipofilling-based reconstruction (LBR) is emerging. In this retrospective study, we compared the surgical and cosmetic outcomes of LBR to combined breast reconstruction (CBR) using fat grafting in conjunction with prosthetic placement.

**Methods:** This single-center retrospective cohort study included 170 patients who underwent mastectomy between 2014 and 2018 and received either LBR or CBR. The surgical parameters and outcomes were compared between the two groups.

Additionally, the cosmetic outcome was evaluated using a Likert-type ordinal scale by the patients, surgeon, and nurse.

**Results:** The study included a total of 170 patients, with 91 patients receiving LBR and 79 patients receiving CBR in combination with prosthetic placement. Patients who received exclusive lipofilling had more sessions on average than those who received prosthetic placement (3.7 lipofilling sessions vs. 2.17 sessions). Additionally, the volume of injected fat was significantly higher in the lipofilling group. No reconstruction failures were reported in either group. Patient, surgeon, and nurse satisfaction scores were high, with an average score of 4.7/5, 4.8/5, and 4.8/5, respectively, at 24 months of follow-up.

**Conclusion:** In this follow-up study, we demonstrate that exclusive lipofilling results in satisfactory cosmetic outcomes with a low rate of complications. This minimally invasive approach to breast reconstruction can serve as a viable alternative to flap and prosthetic-based reconstruction.

## INTRODUCTION

Delayed breast reconstruction (DBR) following total mastectomy, with or without radiotherapy, typically involves free or pedicled autologous flap procedures.

However, these approaches may not be possible or desirable for some patients due to concerns about additional scars or invasive surgeries. In such cases, prosthetic-based reconstruction may be considered as an alternative. Unfortunately, prosthetic breast reconstruction after radiation is associated with a high rate of complications and often leads to poor cosmetic outcomes. 1-3.

Since Coleman’s work in the early 1990s, autologous fat transfer (AFT) has become an increasingly popular approach for managing cosmetic sequelae after breast cancer surgery, particularly in the context of secondary breast reconstructions 4-7. Recent studies have shown that pre-pectoral autologous fat transfer prior to prosthesis placement can improve skin trophicity and vascularization, leading to better cosmetic outcomes. AFT allows for defect areas to be filled, improves overall shape and outline, and increases skin flexibility 7. Preliminary evaluations of this approach have shown promising results, with reduced complications associated with prosthetic reconstruction when lipofilling is used 8,9.

A recent meta-analysis of 21 studies, analyzing 1,011 lipofilling-based breast reconstructions in 834 patients, found that between 2.84-4.66 sessions were required to complete the reconstruction 10. Irradiated patients required significantly more fat grafting sessions to complete the reconstruction compared to non-irradiated patients (4.27 vs. 2.84; p < 0.05). Complication rates were mainly related to radiation therapy, with a rate of 5.4% in the irradiated group compared to 1.1% in the non-irradiated group (considering only necrosis and ulceration). These findings represent a significant improvement from the high rate of complications associated with prosthetic reconstruction without lipofilling in irradiated patients. Additionally, our group has previously reported the efficacy of lipofilling in combination with prosthetic reconstruction in several studies.

Recently, we have experienced an increase in demand from patients seeking to avoid prosthetic placement. In response, we have proposed exclusive lipofilling-based reconstruction as an alternative. In this study, we compared patients undergoing exclusive lipofilling-based reconstruction (LBR) to those having a combined approach of lipofilling and prosthesis (CBR). Our findings showed that both techniques yielded similar results in terms of volume and patient satisfaction. While exclusive lipofilling required more sessions, it was a safe and viable option for patients who preferred not to have a prosthesis.

## PATIENTS AND METHODS

### Patients and procedures

We conducted a retrospective study to evaluate our routine clinical practice between 2014 and 2018 at the Nice Breast Institute. The study was reviewed and approved by the Weill Cornell Medicine in Qatar IRB (IRB19-00155). Our study included only patients who underwent modified radical mastectomy, with or without external chest wall radiotherapy. All reconstructive procedures were discussed and validated in a multidisciplinary tumor board prior to surgery and performed as routine clinical care. The type of reconstruction was chosen based on the patient’s preference and the surgeon’s assessment of skin condition, including factors such as flexibility, pre-thoracic thickness, trophicity, and range of motion of the upper limb.

### All procedures were carried as described below

The first step of the reconstruction consisted of chest wall lipofilling, performed in an outpatient setting at least three months after the end of radiotherapy. Fat was aspirated using a 3 mm liposuction cannula, connected to a 600 ml Redon vial (FMM, France), and a liposuction device (Liposurg, Nouvag, France). The fat was then centrifuged for 30 seconds at 3000 RPM and injected through multiple passages in different planes using a 10 mm syringe and a 1.6 mm cannula. Radial and retrograde injections were used, and percutaneous rigotomy maneuvers were performed when required to remove adhesions between the skin and the deeper plane. The volume of fat injected was distributed as close as possible to the natural shape of the patient’s breast, with particular attention given to defining the infra-mammary fold. No abdominal flap or stitches were usedAll patients were reviewed 4 weeks and 3 months after lipofilling in order to evaluate the skin condition (thickness, flexibility and laxity) and the surgeon decided on one of the following three options:

a. Further lipofilling procedure if the chest wall was not thick and mobile enough for a breast implant,
b. Implantation of a silicone prosthesis if the lipofilling resulted in optimal skin trophicity, thickness and mobility.
c. Finalization of reconstruction with lipofilling when skin condition was considered correct. Volume was determined by patients’ choice.

Second step: implant placement

Patients were assessed for skin trophicity, thickness and underwent up to 3 lipofilling sessions before implant placement if required.

Round implants were used based on the width, height, and projection of the breast to be reconstructed. Incisions were systematically made in the mastectomy scar at its external part to avoid creating new scars. Pre-pectoral dissection was carried out 1cm under the horizontal tangent passing through the infra-mammary fold of the contralateral breast to ensure optimal placement.

Patients who did not wish to undergo implant-based reconstruction underwent exclusive lipofilling-based reconstruction. The number of surgeries required was determined based on the volume needed to achieve the patient’s desired outcome. No drain was placed during the surgery. Following the surgeries, patients received daily assessments from a nurse for a week to monitor the dressing of scars. Patients also wore a compression garment over the liposuction areas for 15 days. Cosmetic results were evaluated by patients, surgeons, and nurses using a Likert-type ordinal scale ranging from 1 (very disappointed) to 5 (very satisfied).

### Statistical analysis

All quantitative data were expressed as mean ± standard error of the mean (SEM). Statistical analysis was performed by using SigmaPlot 11 (Systat Software Inc., Chicago, IL). A Shapiro-Wilk normality test, with a p=0.05 rejection value, was used to test normal distribution of data prior to further analysis. All pairwise multiple comparisons were performed by one way ANOVA followed by Holm-Sidak posthoc tests for data with normal distribution or by Kruskal-Wallis analysis of variance on ranks followed by Tukey posthoc tests, in case of failed normality test. Paired comparisons were performed by Student’s t-tests or by Mann-Whitney rank sum tests in case of unequal variance or failed normality test. Statistical significance was accepted for p < 0.05 (*).

## RESULTS

Between January 2014 and December 2018, a total of 170 patients underwent either exclusive lipofilling-based reconstruction (LBR) or combined lipofilling and prosthesis-based reconstruction (CBR). The demographic characteristics of the patients were consistent with previously described cohorts in the literature (Table 1). The overall average age of the patients was 54.4 years (range: 32-80 years), with patients in the exclusive lipofilling group being significantly older than those in the combined reconstruction group (56.6 vs. 53.3 years). The mean BMI was 23.6 (range: 22-30) and did not differ significantly between the two groups. Of the 170 patients, 126 had secondary breast reconstruction, 9 had prosthesis removal, and 35 had immediate breast reconstruction. A total of 135 patients had received radiation therapy, with no significant difference between the two groups. The average time between the end of radiation therapy and the first lipofilling session was 16.9 months (range: 3-120 months) and was similar between the two groups.

**Table 1.**
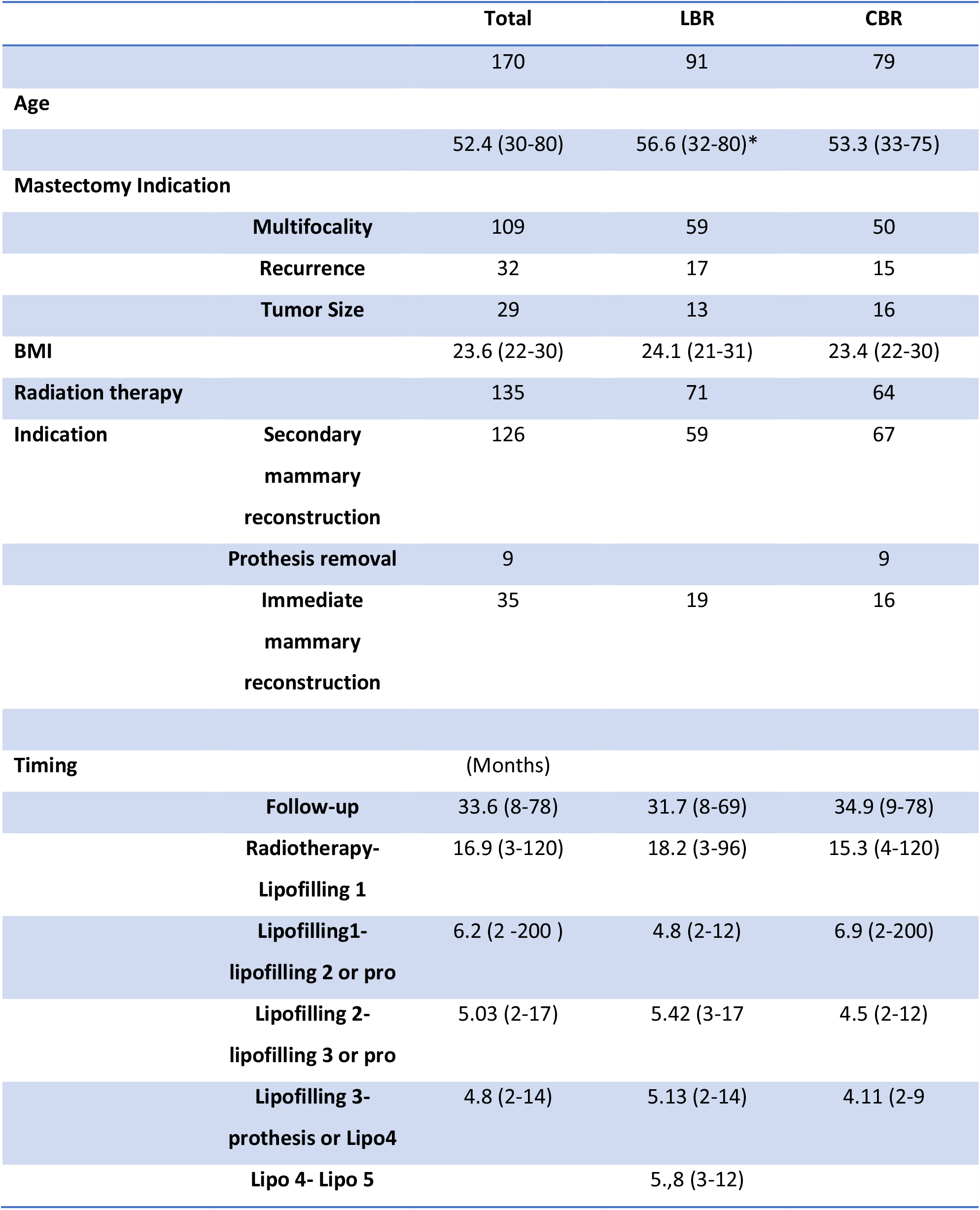
Demographic and pre-operative characteristics of the patient

A total of 59 patients underwent early reconstruction, with the procedure initiated between 3 to 6 months after radiation or mastectomy. We found no correlation between the delay before the first lipofilling session and the number of lipofilling sessions, final cup size, or complication rate. The average delay between lipofilling sessions was similar between both groups and ranged from 4.1 to 6.9 months (Table 1).

Patients in the LBR group underwent more lipofilling sessions than those in the CBR group, with an average of 3.7 sessions compared to 2.17 sessions, respectively (see Table 2 for details). The average volume of injected fat during the first lipofilling was 287 ml (range: 100-600 ml), with no significant differences between the groups.

**Table 2.** Operative characteristics of the patients

|  | **Total** | **LBR** | **CBR** |
| --- | --- | --- | --- |
|  | 170 | 91 | 79 |
| **Volume injected** |  |  |  |
| **Total** |  | 663 ( 240-1010)* | 318 (150- 830) |
| **Lipofilling 1** | 144cc (40-400) | 148 (50-400) | 139 (40-290) |
| **Lipofilling 2** | 165 (60-300) | 175 (80-300)* | 153 (60-280) |
| **Lipofilling 3** | 173 (90-315) | 185 (90-315)* | 145 (90-262) |
| **Lipofilling 4** | 174 (80-320) | 179 (86-320) | 159 (80-280) |
| **Lipofilling 5** |  | 120 (60-250) |  |
| **Prothesis volume** |  |  |  |
| **1 lipofilling** |  |  | 375 ml (190-550) |
| **2 lipofillings** |  |  | 368 ml (275-620) |
| **3 lipofillings** |  |  | 375 ml (175-490) |
| **4 lipofillings** |  |  | 296.7 (150-350) |
| **Cup Size** |  |  |  |
| **B** | 37 | 26* | 14 |
| **C** | 121 | 62 | 56 |
| **D** | 12 | 3 | 9* |

However, patients with exclusive lipofilling had significantly more fat injected at sessions 2 and 3 (Table 2). The volume of the third and fourth lipofilling was also significantly higher than the first and second lipofilling for the LBR group (p<0.05). The total volume of fat injected was significantly higher in patients with LBR (663 +/-175 ml) compared to CBR (320 +/-158 ml) (p=0.002).

Seven patients developed a cystic seroma after the first and second lipofilling sessions; however, reconstruction was carried out as usual, and the cystic seroma resolved during subsequent lipofilling sessions. The mean volume of the prosthesis used in the CBR group was 366 ml (range: 160-770 ml). The mean volume of the prosthesis used was 361 ml (range: 195-525 ml) for one lipofilling, 373 ml (range: 290-620 ml) for two lipofillings, and 381 ml (range: 150-490 ml) for three lipofillings. Patients who underwent four lipofilling sessions had a lower prosthesis volume of 296.7 ml (range: 150-350 ml).There were significantly more patients with a B cup size in the LBR group and significantly more patients with a D cup in the CBR group.

The cosmetic results were evaluated by patients at the end of the procedures, and there were no significant differences between LBR and CBR in terms of outcome, with an average score of 4.8 and 4.7, respectively. Interestingly, we observed that patients were either very satisfied with a score of 5 or unsatisfied with a score of 3. Unsatisfactory results were primarily due to capsular contraction or lack of symmetry, as displayed in Figure 5. The initial cosmetic results appeared to be stable over time, with a follow-up period of 22.7 months (range: 8-45 months). In the CBR group, 9% of patients developed capsular contracture.

## DISCUSSION

In this study, we compared patients who underwent exclusive lipofilling breast reconstruction to those who underwent combined breast reconstruction using both lipofilling and prosthetic reconstruction and found that both groups achieved similar outcomes with high patient satisfaction. Patients who underwent LBR required more sessions and resulted in higher volumes of fat being injected, while the CBR group had higher cup sizes overall. However, high cup sizes were also achievable for patients undergoing LBR. Our complication rate was low and mainly consisted of cystic seromas, with no significant difference between the groups. All procedures were performed on an outpatient basis, including prosthesis placement. The combination of lipofilling and prosthetic placement can be considered a minimally invasive breast reconstruction protocol, with lipofilling sessions performed without scarring and in an outpatient setting, while prosthetic placement is performed through the mastectomy incision, also in an outpatient setting.

Secondary breast implant reconstruction can have high early and long-term failure rates due to previous radiation therapy or poor skin condition, including retraction and adherence. However, skin preparation using fat injection has been shown to significantly reduce the complication rate. Several studies have reported the successful use of autologous fat transplantation, particularly for patients who have undergone radiation therapy 11-15. The complication rate of lipofilling-based reconstruction appears to be low across different studies, particularly regarding reconstruction failure. AFT significantly improves reconstructive outcomes, enabling prosthetic reconstruction of patients with major skin trophicity issues such as fibrosis, retractions, and adherences. AFT improves skin status and reduces skin damage grades, thus providing a promising option for breast reconstruction. In a systematic review of 292 studies Bonetti et al. demonstrated that hybrid reconstruction using lipofilling resulted in good outcome with low complication rate (7.9%) 16. The safety of such approach in terms of recurrence has also been now widely demonstrated. In their meta-analysis of 558 publications ((1590 and 2657 subjects, respectively, in lipofilling and control groups) showed no difference in events occurrence between the two groups.

Our study found that 95.5% of patients rated their reconstruction as very satisfactory or satisfactory, which is higher than the rate reported in a previous study. Although a Likert scale was used in our study, it may have lacked granularity to precisely characterize cosmetic outcomes. However, we found it useful to have a global scale of satisfaction, as demonstrated by our results. Most patients were quite satisfied with the result, as evidenced by a score of 5. Only a few patients had lower satisfaction scores (3), mainly due to a very poor starting condition, resulting in a difficult reconstruction with an average outcome, as shown in Figure 1.

**Figure 1.**
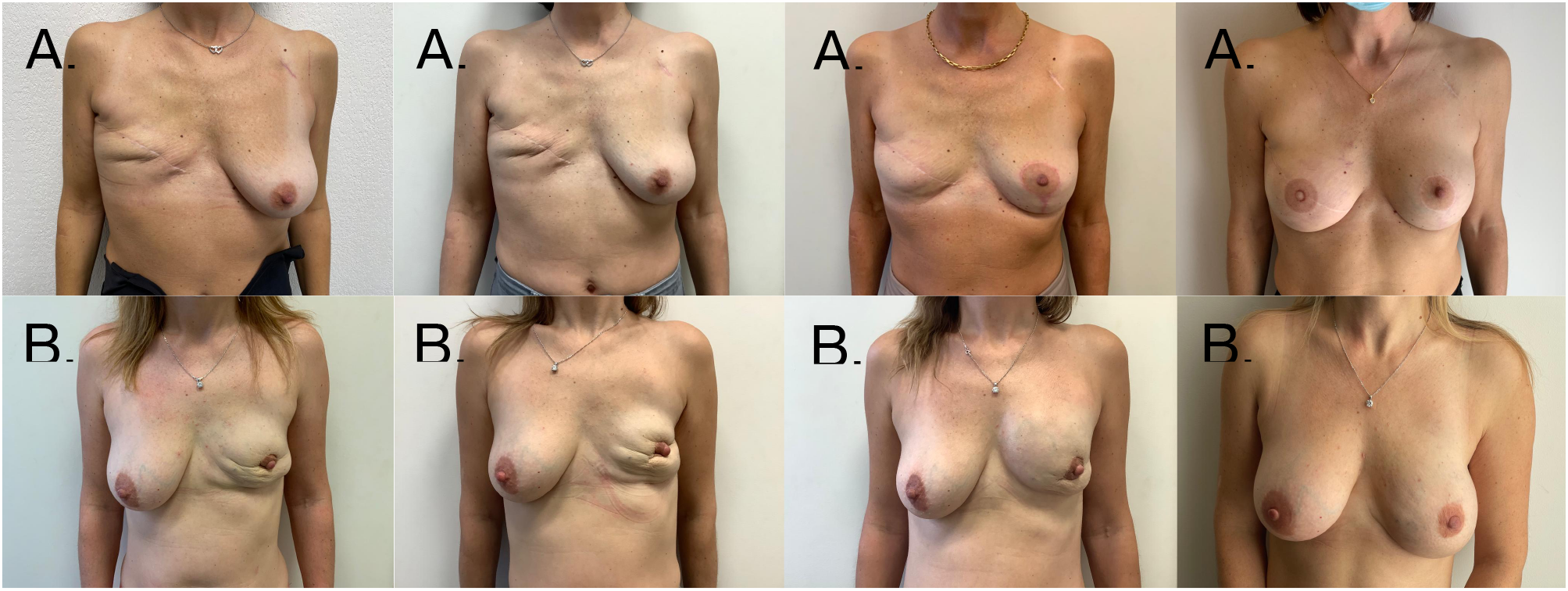
Representative pictures of two patients. Upper patient (A1-4) underwent 3 lipofilling and prothesis placement. Lower patient (B 1-4) underwent prothesis removal and exclusive lipofilling.

This phased approach allows for a gradual and natural-looking reconstruction, while minimizing the risk of complications. It also allows for adjustments to be made based on patient preference and response to treatment. Our study found that patients undergoing LBR required more sessions than those undergoing CBR, but still achieved comparable outcomes in terms of volume and patient satisfaction.

In conclusion, our study supports the use of exclusive lipofilling as a safe and effective option for breast reconstruction, particularly for patients who do not wish to undergo prosthesis placement. Our protocol, consisting of a phased approach, allows for a gradual and natural-looking reconstruction with minimal scarring and low complication rates. Further studies are needed to confirm our findings and assess the long-term outcomes of this approach. Advances in bioreactors and stem cell research may eventually lead to a future where harvested fat can be expanded in vitro for multiple lipofilling sessions, resulting in even less invasive breast reconstruction procedures. With numerous studies demonstrating the positive outcomes of autologous fat transfer-based reconstruction, it is likely that Low Impact Breast Reconstruction (LIBRe) will become the standard of care in the future.

In our setting patients’ choice was the primary factor for choosing between LBR and CBR. Currently patients have a tendency to prefer prosthesis free reconstruction. We do extensively inform patients on the need for multiple sessions and in our experience, we have had no complaints about the higher number of sessions required for LBR. Our outpatient surgical setting is quite appropriate with patients being able to resume normal daily activities within few days of their surgery. It is interesting to note that patients with early reconstruction initiation did not display higher rate of complications and had similar surgical process as well as outcome.

Hence early initiation of minimally invasive reconstruction can be safely initiated 3 to 6 months after radiation therapy. Such approach could greatly enhance patients’ quality of life after mastectomy.

## Data Availability

All data produced in the present study are available upon reasonable request to the authors

